# Predicting the combined effects of case isolation, safe funeral practices, and contact tracing during Ebola virus disease outbreaks

**DOI:** 10.1101/2022.10.06.22280767

**Authors:** Aliou Bouba, Kristina Barbara Helle, Kristan Alexander Schneider

## Abstract

**Background:** The recent outbreaks of Ebola virus disease (EVD) in Uganda and the Marburg virus disease in Ghana reflect a persisting threat of *Filoviridae* to the global health community. Characteristic of *Filoviridae* are not just their high case fatality rates, but also that corpses are highly contagious and prone to cause infections in the absence of appropriate precautions. Vaccines against the most virulent Ebolavirus species, the Zaire ebolavirus (ZEBOV) are approved. However, there exists no approved vaccine or treatment against the Sudan ebolavirus (SUDV) which causes the current outbreak of EVD. Hence, the control of the outbreak relies on case isolation, safe funeral practices, and contact tracing. So far, the effectiveness of these control measures was studied only separately by epidemiological models, while the impact of their interaction is unclear.

**Methods and findings:** To sustain decision making in public health-emergency management, we introduce a predictive model to study the interaction of case isolation, safe funeral practices, and contact tracing. The model is a complex extension of an SEIR-type model, and serves as an epidemic preparedness tool. The model considers different phases of the EVD infections, the possibility of infections being treated in isolation (if appropriately diagnosed), in hospital (if not properly diagnosed), or at home (if the infected do not present to hospital for whatever reason). It is assumed that the corpses of those who died in isolation are buried with proper safety measures, while those who die outside isolation might be buried unsafely, such that transmission can occur during the funeral. Furthermore, the contacts of individuals in isolation will be traced. Based on parameter estimates from the scientific literature, the model suggests that proper diagnosis and hence isolation of cases has the highest impact in reducing the size of the outbreak. However, the combination of case isolation and safe funeral practices alone are insufficient to fully contain the epidemic under plausible parameters. This changes if these measures are combined with contact tracing. In addition, shortening the time to successfully trace back contacts contribute substantially to contain the outbreak.

**Conclusions:** In the absence of an approved vaccine and treatment, EVD management by proper and fast diagnostics in combination with epidemic awareness are fundamental. Awareness will particularly facilitate contact tracing and safe funeral practices. Moreover, proper and fast diagnostics are a major determinant of case isolation. The model introduced here is not just applicable to EVD, but also to other hemorrhagic diseases such as the MVD or the Lassa fever.

## Introduction

After three cases of the Marburg virus disease (MVD) in Ghana [1, 2], the recent spread of the Ebola virus disease (EVD) in Uganda [3] marks the second outbreak of a filo virus in Africa in 2022. The Ebolavirus (EBOV) and the Marburgvirus (MARV) are the most prominent genera of the family of *Filoviridae*, which are non-segmented, negative-sense, single-strained RNA viruses [4, 5].

Both EBOV and MARV are highly contagious and lethal pathogens, classified as biosafety level 4 (BSL-4) agents and category A list pathogens [6], causing hemorrhagic fevers in humans and primates. Index cases emerge from zoonotic reservoirs [7]. Although the reservoirs have not been identified with certainty [8, 9], bats are suspected [10–12]. It is believed that EVD mostly spreads to humans by contact with primates, which have been infected through contact with infected bats [13]. Human-to-human transmission occurs by contact with blood and body liquids of symptomatic persons and infected corpses [14]. In particular, corpses of deceased persons are extremely contagious and both EVD and MVD have been reported to spread during unsafe funeral practices [15, 16]. Five EBOV species, four of which are known to cause EVD in humans [17], have been identified with substantially varying contagiousness and case fatality rates (25% to 90%) [18]. The current outbreak in Uganda is due to the *Sudan ebolavirus* (SUDV), which also caused a significant outbreak in Uganda in 2000/2001, and the first recorded EVD outbreak in 1976 in Sudan [17]. Because the international response was slow, the rather distinct and most prominent species, the *Zaire ebolavirus* (ZEBOV), was identified first. The ZEBOV is the most recurrent, contagious, and lethal species and was also the first one to be discovered in 1976 [18]. By far the majority of EVD outbreaks were caused by the ZEBOV followed by the SUDV with two larger and several minor outbreaks; all other EBOV species caused only minor outbreaks [19].

The Ebola epidemics from 2014-2016 in three West African countries (Guinea, Liberia and Sierra Leone) and from 2018-2020 in the Democratic Republic of the Congo (DRC), both caused by the ZEBOV, have been by far the largest recorded EVD outbreaks [4, 20–25], and substantially challenged global health-emergency management [26]. The outbreak from 2014-2016 amounted to 28,600 cases and 11,325 deaths, yielding a case fatality rate of 39.59% [12, 27–29]. During the outbreak from 2018-2020 in two Eastern provinces of DRC 3,453 cases and 2,273 deaths were recorded [24], resulting in a case fatality rate of 67% [22, 27]. The severity of the outbreak in DRC was fuelled by armed conflicts in the affected areas [24, 30]. The government and international community had only limited access to the affected areas, which had only poor-quality health centers, thereby increasing mortality [25, 31].

Regarding the pathogenesis of EVD, the incubation period ranges from 2 to 21 days [32, 33]. (Notably, the same range is commonly reported for the MVD, but was recently found to be an underestimate [2].) EBOV infects many types of body cells, and thereby produces EBOV glycoproteins that attach to the inside of blood vessels, rendering them to be more permeable [5, 34, 35]. The increased permeability causes the blood vessels to leak blood [36]. EBOV also invades other body parts and organs (liver, spleen, kidney, and brain), which can lead to organ failure and death [37]. The virus also counteracts the host’s natural defense system, by infecting immune cells [4]. Although it is unclear whether survival of EVD confers permanent immunity (because this can only be ascertained during large epidemic outbreaks), evidence suggests long-lasting immunity after recovery [38, 39].

The symptomatic phase of EVD is characterized by a sudden rise in temperature, weakness, muscular pain, headache, and pain in the throat during days 1-3 [18]. During days 4-7 cutaneous eruptions, renal and hepatic insufficiency, internal and external bleeding can occur after the appearance of vomiting and diarrhea [40]. Finally, infected individuals may present with confusion and may exhibit signs of internal and/or visible bleeding, potentially progressing towards coma, shock, and death during days 7-10 [41].

So far, no approved drug treatment exists for the EVD. However, some treatments are associated with improved clinical outcomes [42–45]. Re-hydration therapy and infusions are known to reduce the severity of symptoms [46–49]. Since 2017 three vaccines against the ZEBOV species have been approved [25, 50–54]. Pre- and post-exposure vaccination was a cornerstone of disease management during the 2018-2020 EVD outbreak in DRC [55]. For the recent outbreak of the SUDV in Uganda, the efficiency of current vaccines is unclear [56, 57], but it is assumed that the current vaccines are ineffective [58].

Contact tracing and quarantine strategies are the most important pillars of managing EVD outbreaks [59, 60]. Moreover, safe funeral practices are fundamental [26, 61–63]. These are challenges in the beginning of an outbreak, because of a lack of on-site infrastructure to diagnose *Filoviridae* by PCR [64]. In fact, the index case in the recent MVD outbreak was unsafely buried because the virus was diagnosed *post mortem* [2]. Another problem arose from the underestimated incubation period [65, 66]. Namely, the secondary cases developed symptoms after they completed a 21-days quarantine. Due to the similarities of MVD and EVD, the same challenges also apply to the latter. Furthermore, contact tracing and isolation as primary control strategy present significant logistic and economic strains on the public health systems in low income countries [67].

Here, we introduce a predictive model to study the effect of case isolation, safe funeral practices, and contact tracing during EVD epidemics on disease mortality. To the best of our knowledge, this is the first model of EVD which studies the combined effect of safe funeral practices and contact tracing together. The model is a complex extension of an SEIR-type model (see Figure 1 for an illustration). Model parameters, which have mainly been estimated for the ZEBOV (cf. [68]), are chosen from the literature adjusted such that the dynamics reflect the situation in rural areas in Africa.

**Fig 1.**
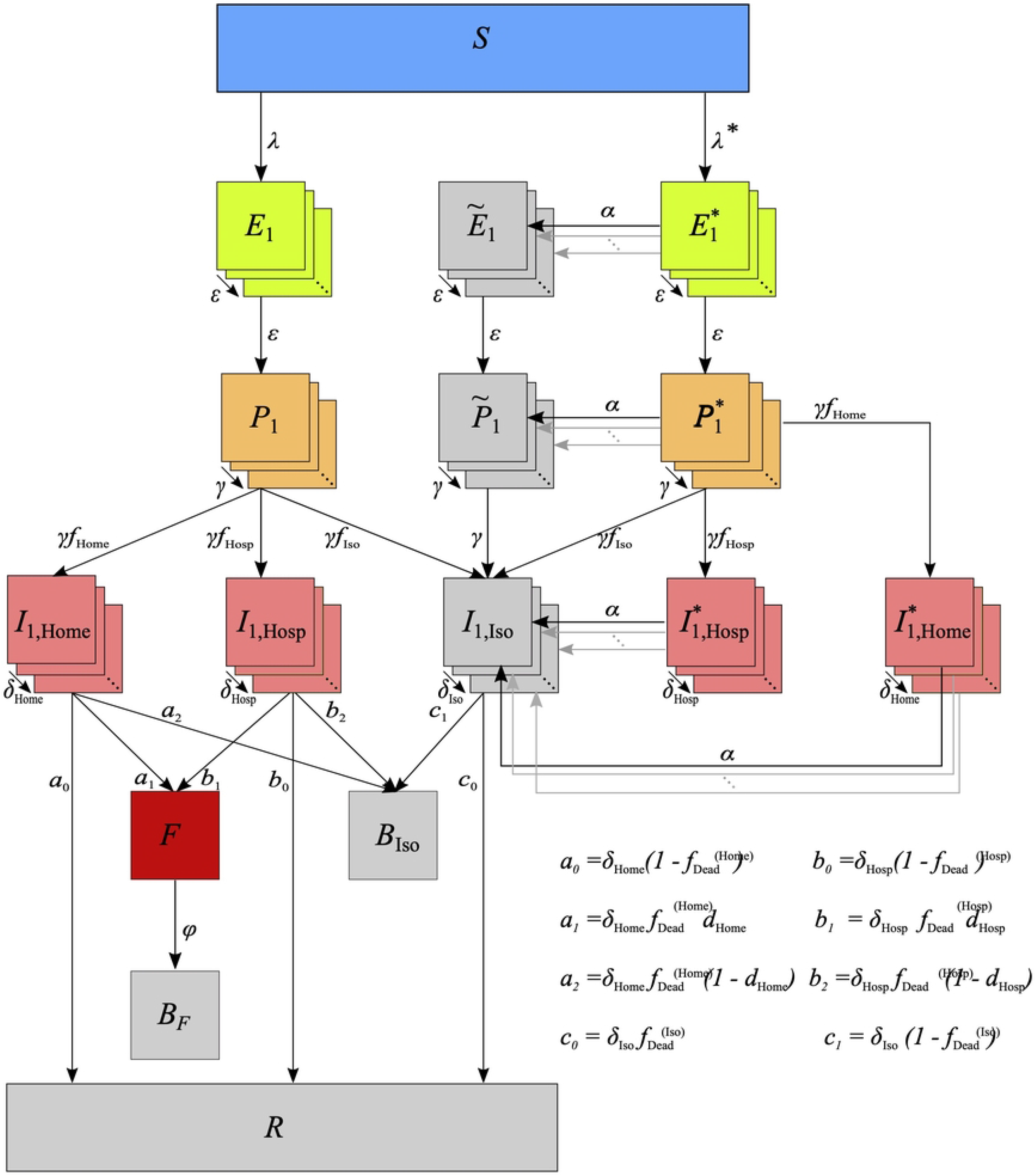
Model. Stages are depicted as boxes, arrows show transition rates. The entire population is grouped into the susceptibles (*S*), the infected which are further classified into the latent (*E*), the prodromal, (*P*), the fully infectious at home, (*I*_Home_), in hospital, (*I*_Hosp_), and in isolation, (*I*_Iso_), and the recovered (*R*). The dead are classified into those awaiting an unsafe Funeral () — still infectious, after funeral (*B*_*F*_), or buried safely (*B*_Iso_). Trace back is modeled through the force of infection for infections subject to trace back *λ*^*^, resulting in individuals in transient stage (*E*^*^, *P*^*^, *I*^(*,Home)^, *I*^(*,Hosp)^) who will get traced back later, and those 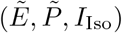 who have already been traced back or diagnosed — these are isolated; *λ* and *E, P, I*_Home_, *I*_Hosp_ describe infections not subject to trace back. The rates *ε, γ, δ*_Home_, *δ*_Hosp_, *δ*_Iso_ describe the progression of the infections. *α* is the rate of successful trace back, *ϕ* the one of funerals, *d*_Home_, *d*_Hosp_ indicate the fractions of safe funerals of non-diagnosed individuals. *f*_Home_, *f*_Hosp_, *f*_Iso_ are the fractions of fully infectious in different treatment, where 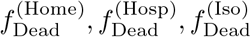 are the related death rates.

The model is *per se* also applicable to the MVD, however, past outbreaks were relatively small compared to EVD outbreaks, so a deterministic approach is questionable for the MVD. In the main text the model is first introduced verbally. A concise mathematical description for readers interested in the technical details is available as Supporting Material. The model is implemented in Python and available at https://github.com/Maths-against-Malaria/Ebola. Outcomes of numerical investigations are presented in the result section.

## Methods

### Basic model compartments

We assume a population of size *N*. First assume no interventions to counteract the EVD outbreak. Susceptibles (*S*) become infected by contacts with infected individuals. Infected first enter the latent phase (*E*), during which they are neither symptomatic nor contagious. This period lasts on average *D*_*E*_ days. The next phase is the prodromal (*P*) phase, which lasts on average *D*_*P*_ days. Prodromals are already infectious, however, not to the full extent, and might develop early symptoms. This phase is followed by the fully infectious phase, during which the disease becomes fully symptomatic. At the end of the prodromal phase, it is determined which percentage of cases will be hospitalized. A fraction *f*_Hosp_ of fully infectious individuals will be hospitalized (*I*_Hosp_), whereas the remaining fraction (*f*_Home_ = 1 − *f*_Hosp_) remains at home (*I*_Home_). Medical treatment is assumed to affect the duration of the fully infectious period. It is assumed that this period lasts an average duration of 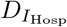 days in hospital, and 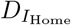 days at home. At the end of the fully infectious period, individuals either recover or die. The fraction of lethal infections at home 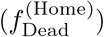 is assumed to be larger than among hospitalized cases 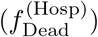. Without proper diagnosis, corpses receive regular funerals (*F*). Importantly, corpses are highly contagious and EVD can spread until the deceased are buried (*B*_*F*_). It takes on average *D*_*F*_ days from death to being buried.

### Adjusting the variance of transition times

An inherent problem with SEIR models are the exponentially distributed transition times from one compartment to the next. Hence, e.g., if the average duration of the latent phase is *D*_*E*_, its variance is 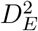. To reduce the variance, instead of modeling the early, prodromal, and fully infectious phases each by a single compartment, they are modeled by several equivalent sub-stages (Erlang-stages), through which infected progress successively (see S1 Appendix for details). Let the number of sub-stages in the respective compartments be denoted by 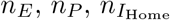, and 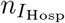. The average duration in the respective sub stages are 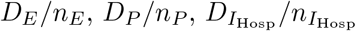, and 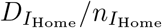. As a consequence, the average duration of the latent, prodromal, and fully infectious phase do not change (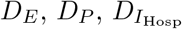, and 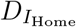). However, the duration are now Erlang distributed and their variances become 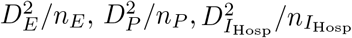, and 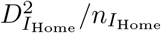. Hence, the variance of the duration can be adjusted by the number of Erlang stages.

### Onset of interventions

To counteract the spread of EVD after its first occurrence at time *t*_Iso_ case isolation, safe funeral practices, and contact tracing are established. To account for these interventions the base model needs to be extended.

### Case isolation

To accommodate case isolation a new compartment for fully infectious but isolated (*I*_Iso_) is introduced, sub-divided into 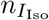 equivalent Erlang stages. It is assumed that, due to precautions, isolated individuals cannot transmit EVD. Since isolated infections are diagnosed, and receive specific medical treatment, the duration during the fully infectious period in isolation is 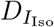 (and 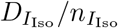 in each corresponding Erlang stage). Also, the fraction of lethal infections in isolation 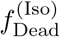 is assumed to be lower than that among hospitalized (and not properly diagnosed) infections.

At the end of the prodromal phase, the proportion of cases *f*_Iso_ which will be isolated is determined. Consequently, the proportions of cases, which will be hospitalized (*f*_Hosp_) and remain at home (*f*_Home_), are adjusted at time *t*_Iso_ to guarantee *f*_Iso_ + *f*_Hosp_ + *f*_Home_ = 1.

### Safe funerals

A characteristic of EVD is that deceased individuals are highly contagious. A recognized concern is the spread of the virus during funeral ceremonies (*F*) [16]. Therefore, all isolated individuals will be buried safely in case of death. To accommodate this, a new compartment of safely buried corpses (*B*_Iso_) is introduced. Transmission does not occur after death if individuals get buried safely. EVD might be properly diagnosed upon death of an individual at home or in hospital, in which case they also receive a safe funeral. It is assumed that fractions *d*_Home_ and *d*_Hosp_ of individuals that died at home or in hospital receive a safe funeral.

### Contact tracing

Contact tracing is a standard practice in many health care systems to contain epidemics. Particularly for diseases as virulent as EVD contact tracing is a cornerstone of disease control. Contact tracing cannot be modeled exactly in an SEIR-type model, since infections are not accounted for on an individual basis. Hence, contact tracing is modeled only approximately.

For contact tracing, infections have to be distinguished into those that are never traced back and not isolated and those that will get isolated and traced back. Contact tracing is not instantaneous, but back-tracking becomes effective after an average duration *D*_*T*_. An individual might be successfully identified by back-tracking during any phase of the disease. To adequately capture this in the model, new compartments for infections (in any of the phases), which will become traced back after an average duration *D*_*T*_ have to be introduced, these are denoted by *E*^*^, *P*^*^, *I*^(*,Home)^, *I*^(*,Hosp)^. All infections, subsumed by these newly introduced compartments, will be isolated before recovery or death. This requires the introduction of new compartments for infections which are isolated after being traced back and isolated (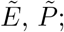 cf. Fig 1). In the fully infectious phase, we no longer have to distinguish between isolated cases which were found by contact tracing and those diagnosed for other reasons (*I*_Iso_). All newly introduced compartments are again modeled by sub-stages. Since the course of the disease is not affected by whether an infections will be diagnosed in the future, the number of sub-stages and the rates of disease progression in the respective phases do not change, i.e., *E*^*^ and 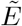 are split into *n*_*E*_, *P*^*^ and 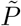 into *n*_*P*_, *I*^(*,Hosp)^ into 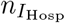, and *I*^(*,Home)^ into 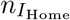 Erlang stages.

### Limited capacity of isolation wards

Infections which are properly diagnosed will be in isolation during the fully infectious phase. Additionally, infections, which were successfully traced back will be isolated during any phase of the infection. Isolated infections are treated in quarantine wards and do no longer contribute to disease transmission. However, it assumed that quarantine wards have a maximum capacity *Q*_max_. The number of isolated cases in excess of this capacity, can no longer be perfectly isolated. It is assumed that only a fraction *p*_Excess_ of these infections is prevented compared to hospital conditions. However, in case an infection that can no longer be properly isolated is lethal, a safe funeral will take place.

### Limited capacity of contact tracing

Due to a lack of capacities, not every individual that should be traced back, can be traced back. There is a maximum capacity *C*_max_ of individuals, whose contacts can be traced back.

### Contact rates

Susceptibles encounter infected individuals (not in isolation) randomly. The relative contagiousness of prodromal individuals, fully infectious individuals at home, fully infectious individuals in hospital, and deceased individuals at unsafe funerals (who are the most contagious) are 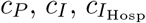, and *c*_*F*_, respectively (typically 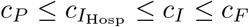). Individuals that will never get traced back and those who will get traced back are equally contagious in the respective disease phases before they get isolated. These parameters affect the contact rates, in the prodromal, fully infectious, and deceased phases, which are denoted by 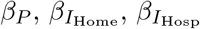, and *β*_*F*_, respectively.

### Implementation of the model

The model as described in S1 Appendix was numerically solved by a 4th order Runge-Kutta method. The code was implemented in Python 3.8 using the function solve_ivp as part of the library Scipy, and library Numpy. Graphical output was created using the library Matplotlib. The implementation of the model can be found at GitHub (https://github.com/Maths-against-Malaria/Ebola).

## Results

The model predictions are first described for a baseline scenario. Subsequently, the effect of (i) case isolation, (ii) safe funeral practices, (iii) contact tracing, and (iv) combined measures and their onset on the peak number of infections and mortality are described. The investigated scenarios differ in their feasibility in terms of logistics, equipment, and human resources. The assumptions range from realistic to ideal.

Importantly, case fatality of EVD varies substantially [18], depending on the viral species. In the main text, we describe the pessimistic situation of high mortality. Additional simulation results assuming moderate mortality are presented in the supplementary figures for comparison. The interpretation is similar to that in the main text.

Model parameters are adjusted to roughly reflect the situation in a rural area (with poor medical equipment) in Africa with a moderate population of *N* = 10, 000. The choices of model parameters are summarized in S2 Table–S3 Table. Initially, (*t* = 0) 10 infections are assumed.

### Baseline scenario

In the absence of interventions, the dynamics follow a standard SEIR model, where an epidemic peak occurs after roughly 200 days with 1,354 active infections (see S7 Table and Fig 2, black line). At this point roughly 44% of the population died or recovered, which coincides closely to the classical “herd-immunity” threshold in SIR models of 1 − 1*/R*_0_ = 0.44 (see Eq. 3.21 in [69]). At this point the epidemic declines and the disease starts to vanish. After roughly 365 days the epidemic is over. The number of remaining susceptibles in the population (2,671) closely resembles the predicted value of 2,675.7 from the final size equation of the standard SIR model (Eq. 1.13 in [69]). The peak number of infections in the latent stage are approximately 700, which are twice those in the prodromal stage (which is intuitive since the latent phase lasts approximately twice as long as the prodromal phase). Roughly 170 cases in the fully infectious phase are hospitalized and the same number remains unhospitalized (this is again intuitive since it is assumed that 50% of cases will be hospitalized). No infection is isolated (Fig 2F) in the baseline scenario and no corpse is buried safely (Fig 2H). More than half of the population dies from the EVD outbreak.

**Fig 2.**
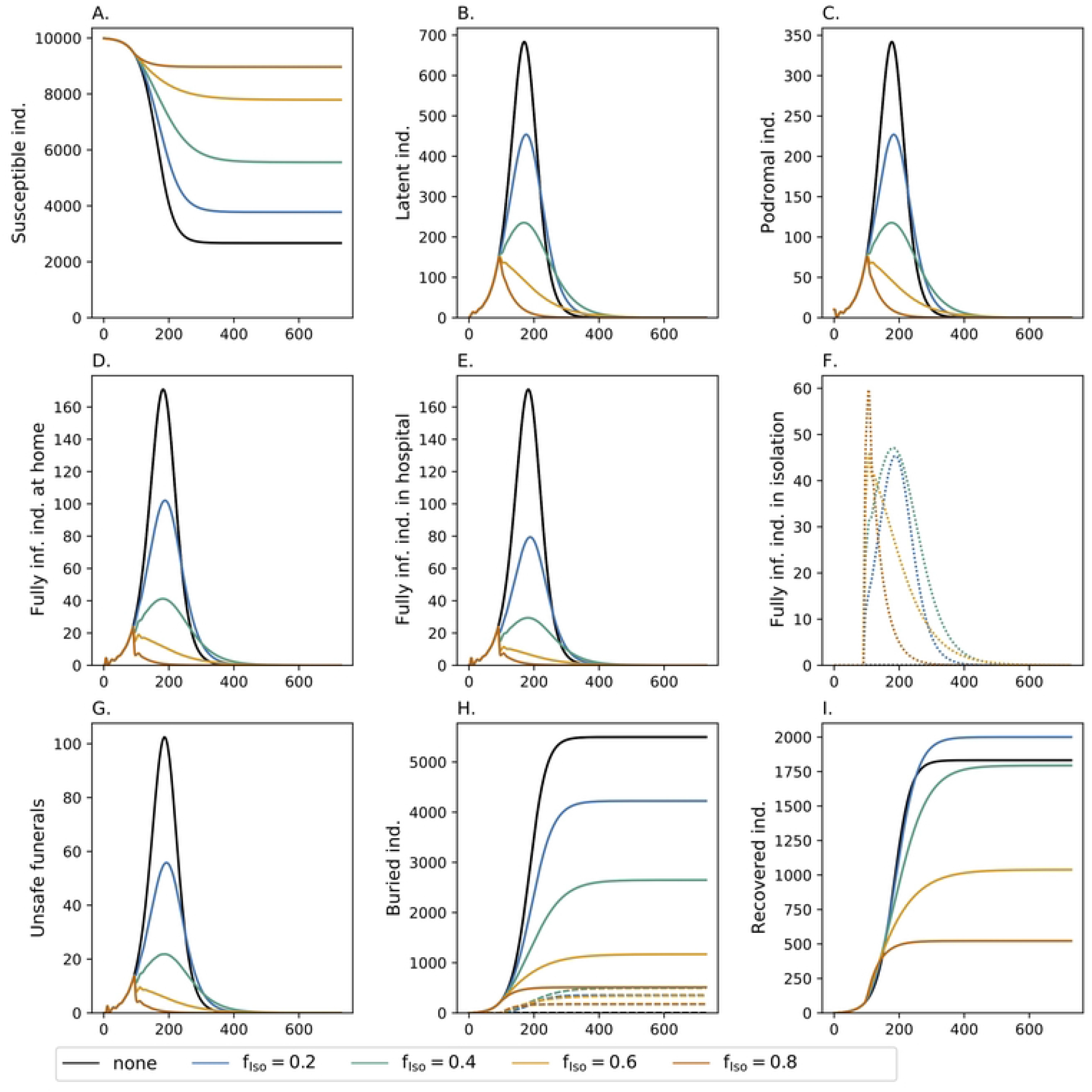
Fraction of isolated *f*_Iso_ infections. Shown are the dynamics of the model for different choices of *f*_Iso_ (colors). Different panels show the number of susceptibles, infected in various phases, unsafe funerals, buried individuals, and recoveries. In the different phases of the infections, the numbers are accumulated over the respective Erlang stages. Isolated infections are shown as dotted lines. In panel (H) the dashed lines show those who have received a safe funeral. Isolation starts at time *t*_Iso_ = 90 days. All other model parameters used for the simulations are listed in Tables S1 Table-S6 Table.

### Case isolation

The effect of case isolation starting at time point *t*_Iso_ = 90 is depicted in Fig 2. Isolating 20% of infections in quarantine wards reduces the slope of new infections. As a consequence, the resulting epidemic peak is lower and occurs slightly later. Hence, also the epidemic ends at a later time point. The overall number of infections is still high with 60% of the population being infected. However, due to isolation, the fraction of infected that remain at home is reduced. Because of better medical treatment in isolation, mortality declines (hence the number of recovered increases in comparison to the baseline scenario) and the number of unsafe funerals decrease by more than 50%. Isolating a larger fraction of infections (40%) has the same qualitative but stronger quantitative effects. Particularly, the epidemic lasts longer, but less than 50% of the population will get infected.

A qualitative change in the dynamics occurs if 60% of cases are isolated. This intervention has an immediate effect. Namely, it readily stops the outbreak with the number of cases starting to decline instantaneously. However, it still takes longer for the epidemic to fade out than in the baseline scenario and overall more than 20% of the population will become infected. Approximately 10% of the population dies from EVD.

The duration of the epidemic is comparable with the baseline scenario if 80% of infections are isolated. In this case around 1,034 individuals become infected and 512 individuals die (S7 Table).

### Safe funerals

Safe funeral practices also help to avoid a large number of infectious contacts, especially because corpses are highly contagious. Safe funeral practices without case isolation (purple line in Fig 3), has approximately the same effect on the epidemic as isolating 20% of the infections (blue line in Fig 2). In combination, safe funeral practices amplify the effect of case isolation. Isolating 20% of the population in combination with safe funerals is comparable to isolating 40% of infections without safe funerals, but mortality is higher. The reason is that case isolation leads to better treatment and hence higher chances of survival. The effect of safe funeral practices in combination with case isolation vanishes if larger fractions of cases are isolated. Namely, isolated cases are always buried safely, and the relative number of deceased that additionally receive a safe funeral decreases. Altogether, this renders isolation and safe funeral practices as insufficient to fully control the epidemic.

**Fig 3.**
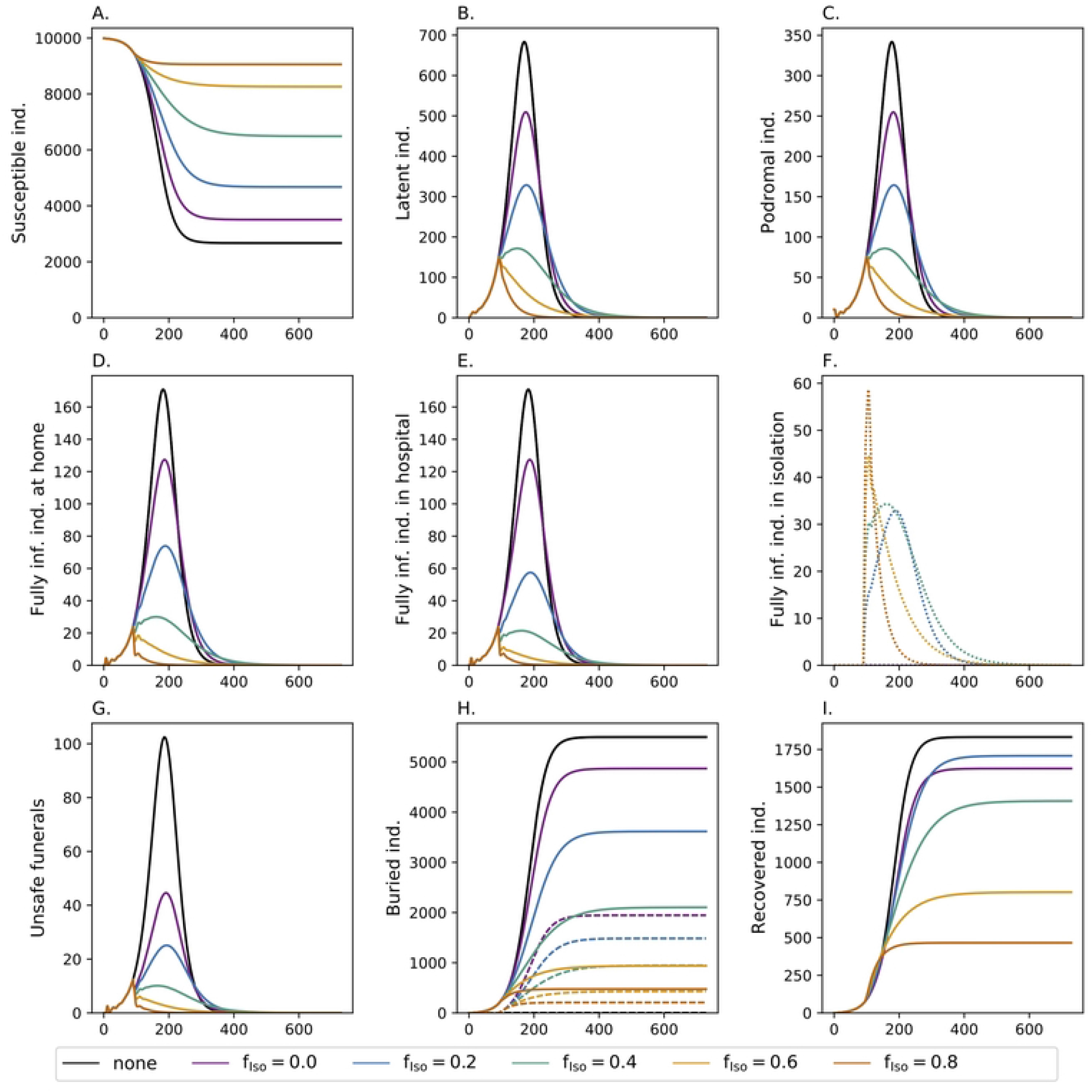
Safe funeral practices. See Fig 2 but combined with safe funeral practices. The fraction of safe funerals after death at home and in hospital are assumed to be *d*_Home_ = 0.16 and *d*_Hosp_ = 0.8, respectively.

### Contact tracing

Tracing the contacts of 80% of the cases that enter isolation can help to identify secondary cases and increase the fraction of infections in isolation (Fig 4). The effects are strongest if isolation by diagnosis (*f*_Iso_) is at 20% to 40%, because many cases will be found and isolated by contact tracing, which would otherwise remain undetected. If initially 80% of infections are diagnosed and isolated, additional isolation by contact tracing has only a small effect. The effects of contact tracing on the epidemic peak and the number of infections are not very strong. However, better treatment in isolation clearly increases the chances of survival. In all scenarios the number of cases in isolation at a time is limited to about 60, which determines the required (effective) quarantine capacity *Q*_max_ (note that in practice also uninfected individuals will get isolated, so the quarantine capacity must be appropriately higher).

**Fig 4.**
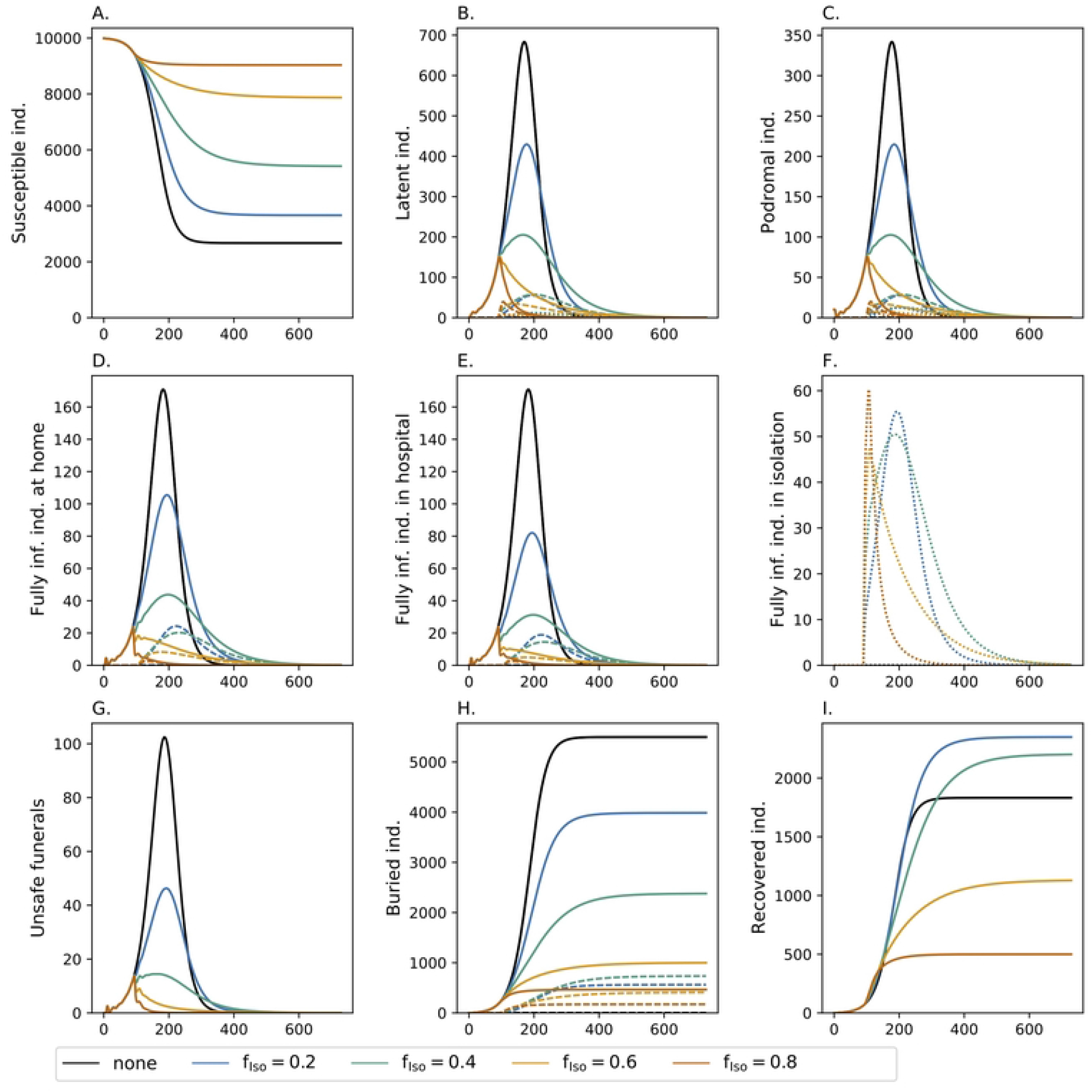
Contact tracing. See Fig 2 but combined with additional contact tracing. A fraction *f*_Tr_ = 0.8 of the contacts of infections in isolation are traced back and isolated themselves. In panels (B-F) the dashed lines show the number of infections that will be traced back at some time in the future (not yet isolated) or are currently traced back (and in isolation). The dotted lines show all individuals currently in isolation. In panel (H) the dashed lines show the numbers of safe funerals that were conducted.

### Extent of contact tracing

The effect of the extend of contact tracing is illustrated in Fig 5 for the ideal case that 80% of infections are isolated (*f*_Iso_) anyway. Tracing 80% of the contacts of isolated individuals only reduced the number of deaths by about 10%.

**Fig 5.**
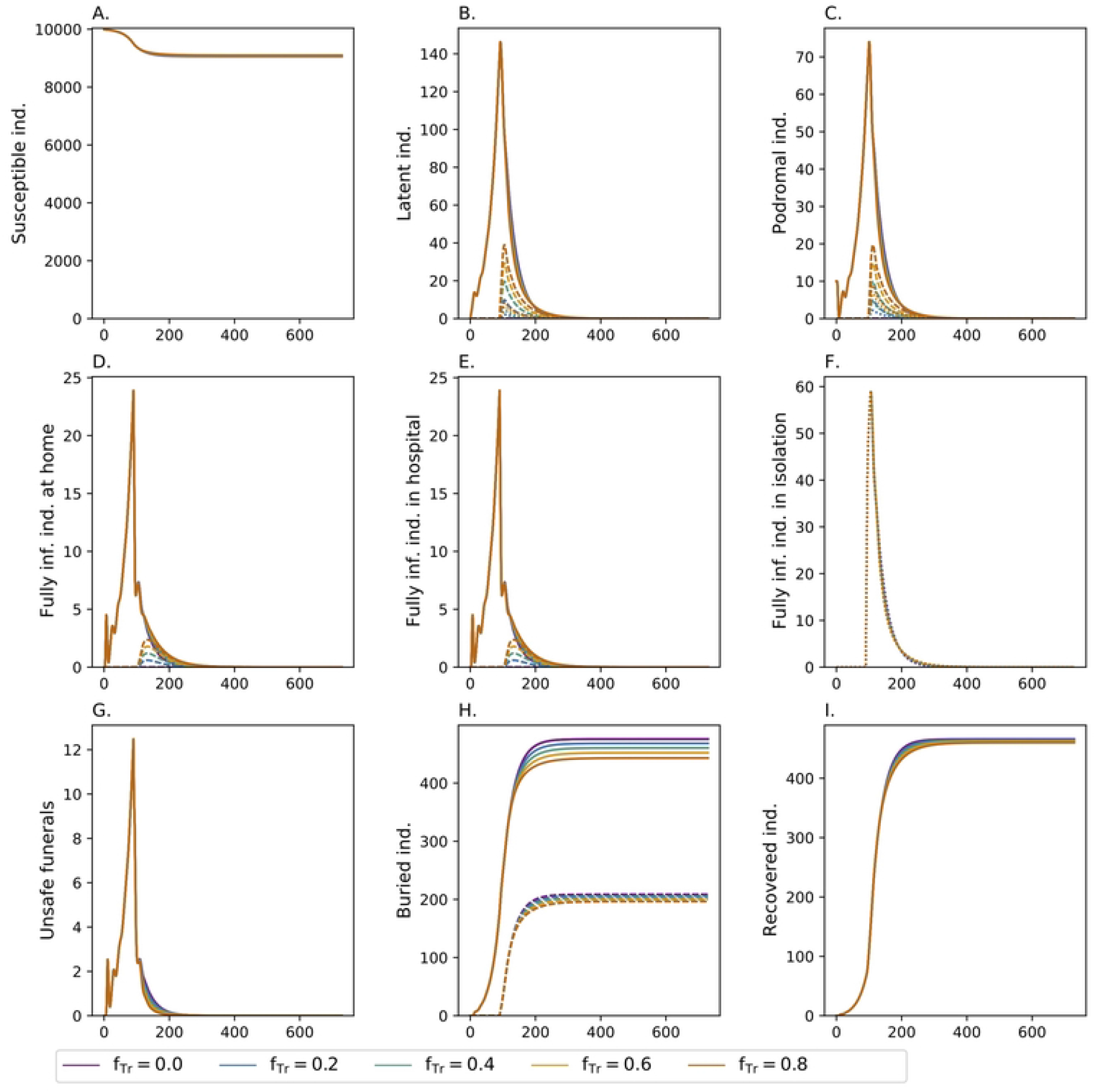
Extent of back-tracking. Effect of fraction of infected individuals who will be traced back *f*_**Tr**_, when the fraction of infections that are isolated is *f*_Iso_ = 0.8 with additional safe funeral practices for lethal cases that occurred outside isolation (*d*_Home_ = 0.16 and *d*_Hosp_ = 0.8). Line types as in Fig 4.

### Efficiency of tracing back

The efficiency of contact tracing can be measured by the average time necessary to isolate suspected infections, i.e., by *D*_*T*_. The effect of the duration necessary to trace back contacts is illustrated in Fig 6. Since it is assumed that cases become fully infectious on average after 15 days, and they recover or die on average 5 days later, shortening the trace-back time from 20 to 5 days, has a substantial effect. This is especially true when reducing the trace-back time from 25 to 10 days. Any further reduction leads only to marginal improvements. The reason is that 20 days are too long to prevent tertiary cases.

**Fig 6.**
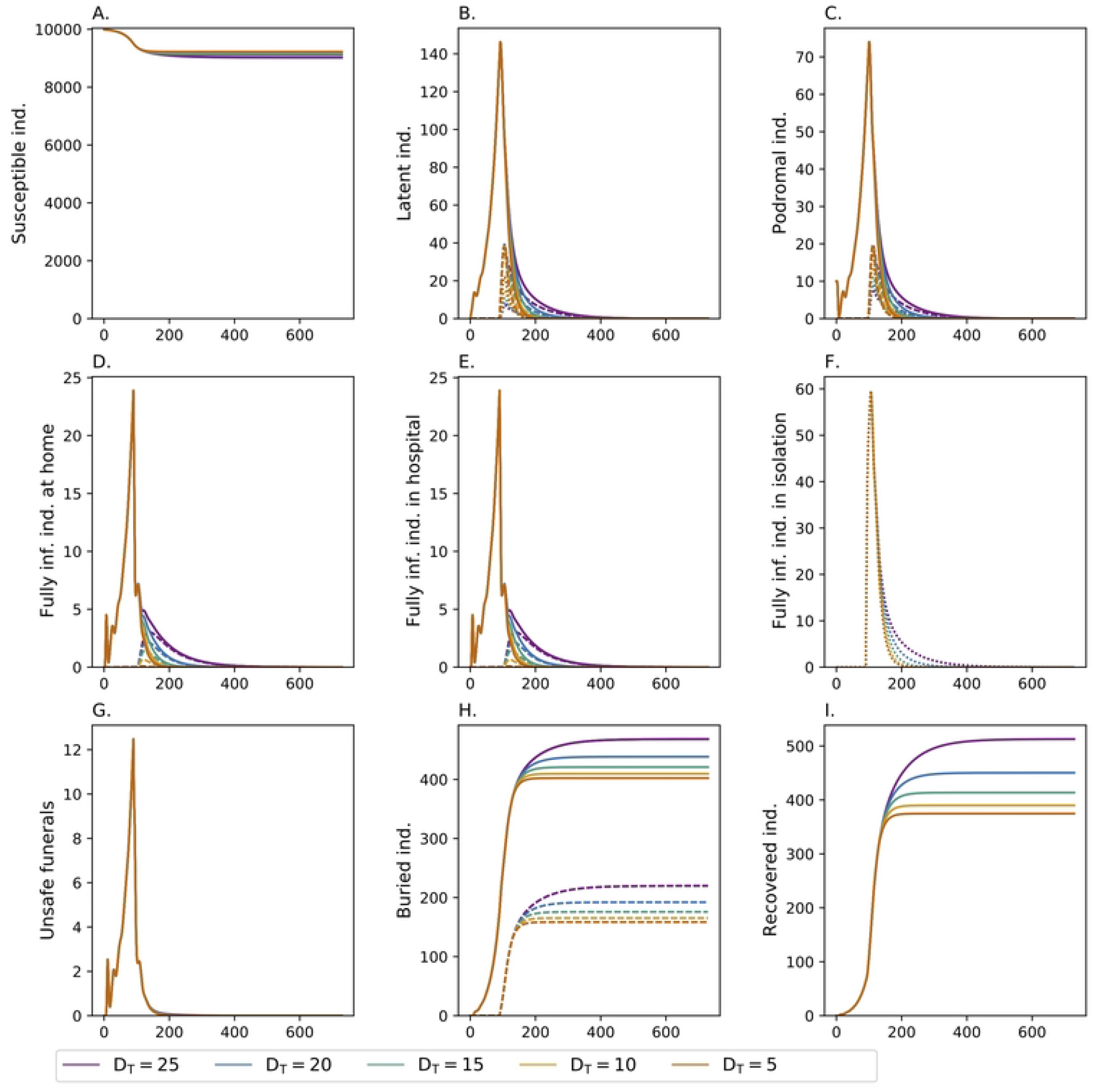
Efficiency of contact tracing: Shown is the effect of the average trace-back time *D*_*T*_ (colors), assuming 80% of infections are isolated (*f*_Iso_ = 0.8) and 80% of contacts of isolated persons are traced back (*f*_Tr_ = 0.8). Additional safe funeral practices for lethal cases that occurred outside isolation (*d*_Home_ = 0.16 and *d*_Hosp_ = 0.8) are assumed. Line types as in Fig 4.

### Combining isolation, contact tracing, and safe funeral practices

As expected, a combining of all three measures leads to the best outcome (Fig 7). The combination of case isolation, safe funeral practices, and back-tracking has a clear effect. Although the effects overlap, e.g., of case isolation and back-tracking, in combination the measures have a synergistic effect. In total, all measures combined compared to just isolation reduces the number of infections and deaths to about 900 and 450, respectively.

**Fig 7.**
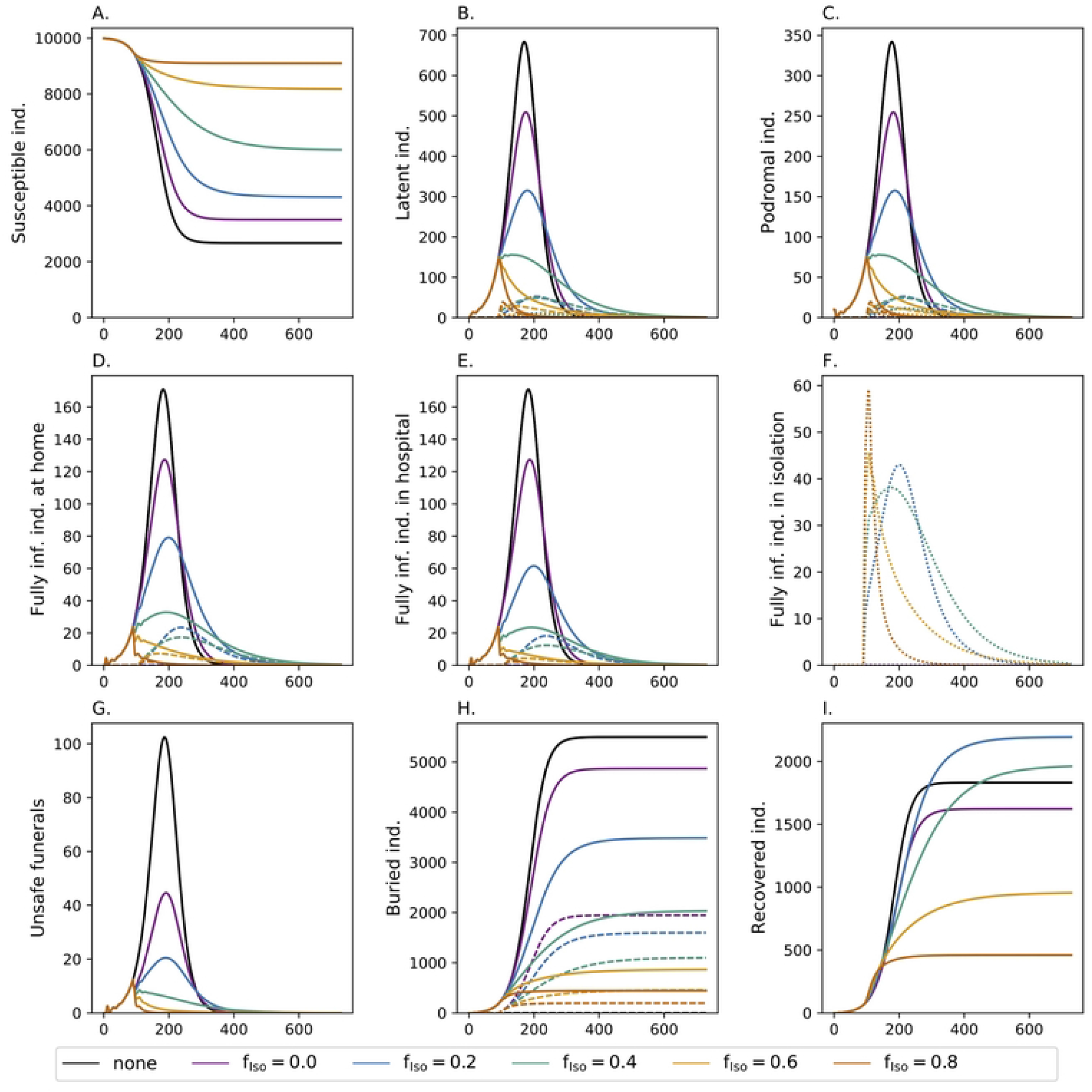
Combination of interventions. See Fig 4 but with additional safe funeral practices for lethal cases that occurred outside isolation (*d*_Home_ = 0.16 and *d*_Hosp_ = 0.8). The average trace-back time is *D*_*T*_ = 21 days. A fraction *f*_Tr_ = 0.8 of contacts of infections in isolation are subject to back-tracking.

### Onset of interventions

The onset of interventions after the first EVD cases started is very important to curtail the spread of the disease (Fig 8, S7 Table col. 5,6). Starting to isolate 80% of cases (*f*_Iso_ = 0.8) and back-tracking 80% of their contacts (*f*_Tr_ = 0.8) *t*_Iso_ = 90 after the occurrence of the index cases in combination with safe funeral practices has already a profound effect in containing the epidemic. However, the earlier these measures are implemented the stronger the effect on the epidemic outbreak. In fact, implementing the measures *t*_Iso_ = 30 days after the index cases occurs, contributes to contain the disease outbreak. In fact, the number of infections is reduced from approximately 900 to 120 and the number of deaths from 450 to 55, respectively.

**Fig 8.**
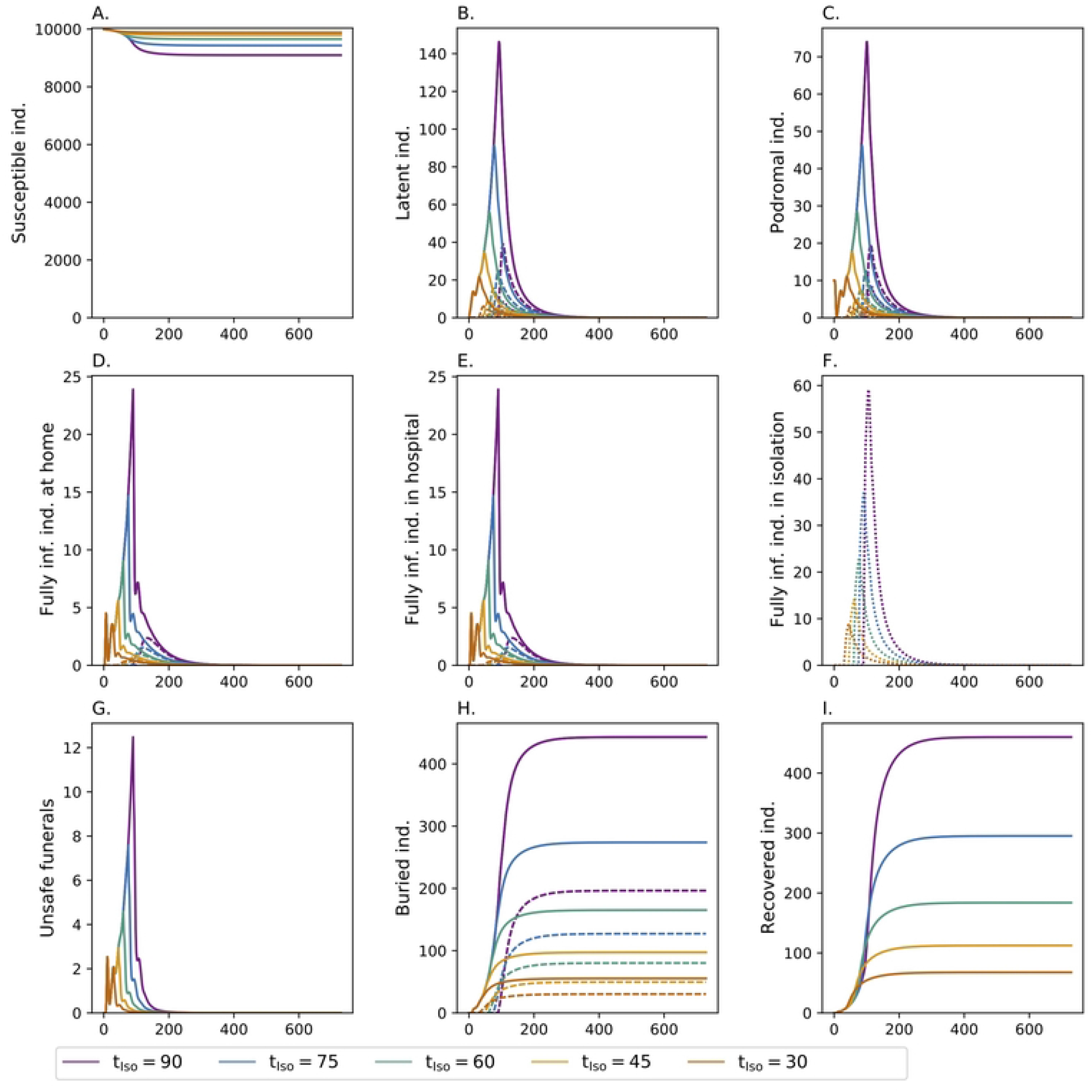
Onset of interventions: Shown is the effect of the onset of interventions *t*_**Iso**_ (colors). The fraction of infections that are isolated is *f*_Iso_ = 0.8, 80% (*f*_Tr_ = 0.8) of the contacts of isolated patients are subject to back-tracking, and safe funeral practices are conducted outside isolation (*d*_Home_ = 0.16 and *d*_Hosp_ = 0.8). Line types as in Fig 4.

## Discussion

The importance of contact tracing in epidemic management is evident in the current outbreak of the Ebolavirus disease (EVD) due to the Sudan ebolavirus (SUDV) in Uganda. Being classified as biosafety class 4 pathogens, immediate action is necessary at each potential outbreak. So far, no effective vaccine against the SUDV exists, unlike for other Ebolavirus species [56–58]. As with all *Filoviridae* no approved treatment exists. Hence, fast and reliable diagnostics, case isolation, safe funeral practices, and contact tracing along with quarantine are the means of containing outbreaks. This also requires a trained force of healthcare workers and appropriate PPE equipment, potentially on a larger scale.

While the appropriateness of the above measures to contain an EVD outbreak is unquestionable, ideally their relative effectiveness in comparison to each other and in combination is quantified, to achieve the optimal response to the outbreak. Clearly, more is better, but given limited resources in terms of qualified personnel, infrastructure, etc., particularly at the onset of an epidemic one should prioritize the most effective measures.

To quantify the effectiveness of several public health responses, we introduced a deterministic, SEIR-type predictive model. The model considers a latent and prodromal phase of the infections. Fully infectious (symptomatic) infections are assumed to be treated at home, in hospital, or in isolation, which reflects the infrastructure of the population (in rural areas it might not be possible/practical to reach a hospital on time, or people avoid hospitals due to widespread superstition etc.) and the capacities to properly diagnose the disease. The fact whether an infection is treated at home, in hospital, or in isolation affects the probability of disease transmission and mortality. Properly diagnosed infections will always be treated in isolation, with appropriate measures [70, 71]. If the disease is not diagnosed properly in hospital, the received treatment might be too unspecific. Importantly, EVD is easily transmitted during unsafe funeral practices. If an infection was treated in isolation or EVD was identified as the cause of death, it is standard practice to bury the corpse safely. Standard protocols typically involve the use of PPE and disinfecting the body bags of the corpse. Furthermore, we included contact tracing in the model, i.e., individuals in contact with infected individuals will be quarantined. The model accounts for the fact that case isolation is not immediate, i.e., it takes time to trace potential contacts.

A common limitation of SEIR-based modes are exponentially-distributed transition times, i.e., the durations of the different phases of the infection follow an exponential distribution. Hence, the average duration already defines its variance, which leads to unrealistic dynamics (cf. e.g. [72–75]). To overcome this problem, we introduced sub-stages of the model compartments (Erlang–stages), which result in Erlang-distributed durations, allowing to adjust their variance.

In the model, it is assumed that contact tracing is not implemented immediately, but only after some delay. The choice of the delay might seem rather long, but should be noted that the disease will only be first reported/discovered after some time delay. Furthermore, the suspicion of EVD must arise. Finally, in a deterministic model with a single index case, some initial time is required to let the number of infections rise to a realistic number. In fact, in the current outbreak, within a relatively short time approximately 30 infections were reported, before contact tracing became effective.

The quality of the medical infrastructure is reflected by the fraction of infections which are isolated. This reflects the availability of hospitals, and resources to properly diagnose EVD. The fraction of individuals, which are isolated, has a substantial effect on the epidemic outcome. Clearly, the more infections are isolated, the lower the epidemic peak and overall mortality.

Also, safe funeral practices (after death at home or in hospital) contribute substantially to contain the epidemic. Namely, corpses are highly contagious and contribute to disease transmission without proper precautions, and safe funeral practices are equivalent to isolation during this phase. If most infections are isolated, a large proportion of infected corpses are buried safely anyway, and the additional safe funerals have only a relatively small effect on reducing the burden of the epidemic. This is hardly surprising, as proper diagnosis occurs already prior to death.

Case isolation and safe funeral practices alone are insufficient to fully contain the epidemic under plausible parameters. This changes if these measures are combined with contact tracing, which additionally contributes to reduce the number of infections and mortality. Importantly, contact tracing and isolation are not independent. Only the contacts of diagnosed infections can be traced, and these will automatically be isolated. Hence, the better the diagnosis (the higher the fraction of isolated infections), the more efficient is the contact tracing.

The quality of contact tracing is not just measured by the fraction of infections, whose contacts will be traced, but also by the average time it takes to trace back the contacts. Shortening the trace-back time has a noticeable effect. Particularly, because the likelihood is larger to isolated contacts, who got infected, before they become infectious.

The model introduced here can be used to explore the optimal strategy of coming up with the above measures. Clearly, the parameters have to be adjusted to a specific situation. However, in areas with suspected zoonotic reservoirs, model parameters can be adjusted to the particular area (in terms of diagnostic, hospital, and logistic capacities) and the optimal response to different Ebolavirus species can be determined as a means of epidemic preparedness. Anyhow, our results suggest that isolation of suspected cases is crucial, implying fast diagnostics and increased awareness in hospitals are fundamental to contain outbreaks. In practice, in the beginning of an outbreak, diagnosis might take substantially long. After suspected cases are reported, public health authorities are informed that involve specialized institutions in the diagnosis.

The model is not just applicable to EVD, but also to other viruses causing hemorrhagic fevers, like the Marburg virus or the Lassa virus. However, the model is deterministic, and thus it is only appropriate for diseases which have a substantially high base reproduction number so that enough cases occur to ignore stochastic effects. This is questionable for the MVD, which had relatively small outbreaks compared to EVD outbreaks.

A downside of the SEIR-based model is that contact tracing can only be captured approximately, because individuals are not modeled explicitly. The logic of the model can be adapted in an individual-based model. However, the deterministic approach allows studying the interaction of control interventions without confounding stochastic factors.

The model assumes that isolation is perpetual until recovery or death, i.e., it is disregarded, that individuals being quarantined leave isolation before the onset of symptoms. This can happen if the incubation period of the EVD is underestimated. In fact, the incubation period was underestimated for the MVD, and in two cases during the outbreak in Ghana the onset of symptoms occurred after the completion of the mandatory quarantine [2]. Given that the incubation period of the EVD was, as most other parameters, mainly estimated for the ZEBOV, it cannot be ruled out that the incubation period of a different EBOV species such as the SUDV actually differs. Importantly, there is definitive evidence of spermatogenic transmission of the MARV [76]. Although, this route of transmission is unclear for EBOV, 12 months of safe sex are recommended after the onset of symptoms [70, 71]. Also, the possibility of spermatogenic infection was not included in the model, however, it should be of limited relevance during larger outbreaks.

The applicability of the model to the ZEBOV, for which vaccines have been approved, is limited. The reason is that pre- and post-exposure vaccination will be pillars of epidemic management of ZEBOV outbreaks, which is not captured by the model. However, the present model serves as a blueprint for further model extensions.

## Data Availability

Only simulated data is reported. The Python code will be made available via GitHub upon publication.

## Acknowledgments

The first ideas of this work were fostered by AIMS (https://aims-cameroon.org/) under the supervision of Martin Eichner (University Tübingen, Germany). His inputs are gratefully acknowledged.

---

**S1 Appendix Mathematical Description**

**S1 Table. Population size and model compartments**.

**S2 Table. Summary of model parameters describing disease progression and choices for the simulations**.

**S3 Table. Summary of model parameters describing infections and choices for the simulations**.

**S4 Table. Summary of possible scenarios once interventions control are set up**.

**S5 Table. Summary of possible scenarios of safe funeral at home and hospital**.

**S6 Table. Summary of possible scenarios (severe and mild mortality) of dead cases at home, in hospital and in isolation**.

**S7 Table. Summary of total infected, maximum infected, and total fatalities**.

